# Does community-based health insurance affect lifestyle and timing of treatment seeking behavior? Evidence from Ethiopia

**DOI:** 10.1101/2023.12.15.23300041

**Authors:** Zecharias Fetene Anteneh, Anagaw D. Mebratie, Zemzem Shigute, Getnet Alemu, Arjun S. Bedi

## Abstract

There has been a growing concern about the financial sustainability of community-based health insurance (CBHI) schemes in developing countries recently. However, little empirical studies have been conducted to identify potential contributors including ex-ante and ex-post moral hazards. We respond to this concern by investigating the effects of being insured on household lifestyle -Preventive Care- and the timing of treatment seeking behavior in the context of Ethiopia’s CBHI scheme. Using three rounds of household panel data and a fixed-effects household model, we do not find a significant impact on preventive care activities. However, we find that participation in CBHI increases delay in treatment-seeking behavior for malaria, tetanus, and tuberculosis symptoms. This behavior is costly for the insurer. Therefore, it is essential to identify the primary causes of delays in seeking medical services and implement appropriate interventions aimed at encouraging insured individuals to seek early medical attention when signs of diseases emerge.

## Introduction

High costs of care in developing countries are recognized as a leading cause of poor health care utilization, poor health outcomes and a key factor in precipitating poverty. According to (1), 800 million people worldwide spend at least 10 percent of their household budget on health care expenses. For nearly 100 million people, that spending is enough to drive them into extreme poverty and force them to survive on just $1.90 or less a day. With the aim of ensuring all people can access the health services they need without facing financial hardships, CBHI schemes have been introduced since the second half of the 19th century in the developing world (2,3). CBHI scheme is typically used to reduce inequalities in health care utilization by reducing the cost of care. This scheme are established on the basis of existing principles of social solidarity, operate at the community level and allow community involvement in scheme management (4). Typically, membership fees are low and substantial portion of healthcare costs are, therefore, subsidized by the government. The scheme differs from social health insurances, which is primarily for employed individuals, and publicly funded health insurance programs, which are essentially free at point of use and tax funded.

Extensive studies evaluating CBHI have been conducted in different countries, mainly examining the impact on health care utilization and out-of-pocket spending (5–12). While CBHI seems promising in terms of enhancing health care utilization, concerns have been raised about its sustainability as a health financing system (13–15). In the context of Tanzania, (16) identified the presence of large claims, leading to the suggestion that policymakers should confront a tradeoff between reducing the health benefit package or increasing revenues to facilitate the nationwide expansion of the existing insurance scheme. This concern resonates with findings from (14), who observed excess claim costs and negative net income within Ethiopian CBHI in specific districts. A similar concern has been expressed by the relevant government officials regarding Ethiopian CBHI (17). Large claims in CBHI may stem from various factors, including ex-ante and ex-post moral hazard problems (18). Ex-post moral hazard refers to the situation where beneficiaries might become excessive healthcare consumers once they are insured, especially those who enroll with severe existing health conditions, driven by a perceived lack of financial barriers to seeking care. They may be more likely to seek medical care, driving up claim costs. On the other hand, ex-ante moral hazard may manifest as a lack of preventive practices or unhealthy behaviors among beneficiaries, leading to a higher likelihood of future health issues and, subsequently, large claims.

While there is limited empirical evidence on whether adverse behavior by beneficiaries contributes to rising healthcare costs in CBHI schemes, studies based on different insurance schemes provide mixed insights. In developed countries, (19–22) suggests that insurance can drive unhealthy lifestyles among policyholders, given that the health consequences of their behavior are shared between insurers and policyholders. Conversely, studies like (18,23–25) report increased utilization of preventive care with no evidence of risky health behaviors among the insured individuals. One perspective on this phenomenon, as discussed in (18), is that while insurance covers the financial aspect of healthcare access, risk-averse individuals might still experience utility losses due to ill health. Consequently, this factor could potentially mitigate the incentives for insured individuals to reduce their preventive efforts.

In the context of low-income settings, (26–28) have made initial attempts to understand behavioral responses to insurance schemes. (27) examined the impact of a CBHI scheme in Burkina Faso on the probability of individuals seeking care when sick while, (26,28) focused on the effects of insurance on preventive activities, such as use of malaria bed nets.

In this paper we examine the effects of Ethiopian CBHI on behavioral response by assessing preventive care activities, such as hand-washing before meals and water treatment before drinking. Additionally, we analyze the timing of care-seeking behavior in hypothetical cases of Malaria, Tetanus, and Tuberculosis symptoms. Our main departure from the literature is while previous studies, such as (12,27,29,30), have primarily focused on the treatment methods chosen by beneficiaries, our study looks at when households decide to seek care upon the onset of disease symptoms. This approach is particularly relevant for policymakers, as delayed care-seeking behavior among beneficiaries can have cost implications for the insurer and potentially impact the financial sustainability of the CBHI scheme.

In Ethiopia, CBHI has been implemented since 2011 to cover people operating in the informal sector (rural and urban). The scheme provides financial protection for low-income families to foster healthcare utilization thereby to improve their health status. Initially, four regions out of nine administrative regions were covered by the program. Within these four regions 13 districts were selected to gauge the response of households and later expanded to cover other districts. Since this pilot stage, by June 2022, 879 districts located in six regions and two city administrations (Addis Ababa and Dire Dawa) have joined the scheme and it has enrolled more than 40% of the population in the country. As part of the Health Sector Transformation Plan, it is also planned to cover 80% of districts and to enroll 80% of the households at national level (Data from EHIS 2022).

Using three rounds of household panel data and applying a household fixed-effect approach, we did not find a significant effect on preventive care behaviors. However, participation in CBHI appears to be associated with increased delays in accessing modern healthcare.

This paper is organized in six sections: section 2 describes Community-Based health insurance, section 3 presents data, section 4 empirical approach, section 5 provides results and discussions, and section 6 concludes.

### Community-based health insurance in Ethiopia

Geographically, Ethiopia is divided into twelve regions and two administrative cities. Within the public health system, the provision of healthcare infrastructure increased between 2000 and 2015 in all parts of the country (8). Following the expansion of healthcare infrastructure, a key motivation was to enhance the accessibility of healthcare services with the assumption that improved accessibility would encourage greater utilization of care. However, despite these efforts, a significant hurdle persisted – individuals were required to cover healthcare expenses out-of-pocket. This financial barrier had a notable impact on healthcare utilization, resulting in continued low rates of utilization. For instance, in the years 2009, 2010, and 2011, outpatient attendance per capita, defined as the total outpatient attendance over the total population, stood at rates of 0.29, 0.3, and 0.29, respectively (31).

In response to this, the CBHI scheme was introduced in June 2011, specifically targeting those engaged in the informal sector both in rural and urban areas. The overall goal of the scheme is to achieve universal health coverage by ensuring low-cost access to health services, especially for those who cannot afford to use them. Initially, the scheme was piloted in thirteen woredas (hereafter districts) selected from four main regions (Tigray, Amhara, Oromia, and SNNP). The complete set of district selection criteria included (1) Willingness of district authorities to implement the schemes (2) Commitment of districts to support schemes, (3) Geographical accessibility of health centers (4) Quality of health centers, (5) The implementation of cost recovery, local revenue retention, and public pharmacy policies in health centers (8). Village level implementation of the scheme is decided by a majority vote of the residing households while households’ enrolment in the scheme is voluntary. To curtail the problem of adverse selection, registration is accepted at the household level. Therefore, all household members must present their family insurance card to access the benefit packages at health care points.

Premium is a fixed rate, unlike other types of insurance that tend to be progressive based on income. This is decided by the federal government in collaboration with local governments and other stakeholders. At the beginning, the annual cost of premiums for households was low, ranging from 126 to 180 Ethiopian Birr (ETB), but from 2020 the premium increased to 350 ($10) for urban households and 240 ETB ($7) for households living in rural areas. This cost amount to 0.5 to 1% of household monthly income (32), but it might vary slightly across regions depending on household characteristics (8). For instance, for a core household with a parent and all children under 18, the monthly premium for the first year is 10.50 ETB in SNNP, 11 ETB in Tigray, and 15 ETB in Oromia. Households pay a one-time registration fee and premiums to the village office, which then transfers the funds to the CBHI scheme (32). The scheme also targets and provides fee waivers to the poorest households in the pilot districts. However, only few of eligible households have actually joined the scheme with fee waiver benefits, not more than 10% of the population in the district.

CBHI benefit package includes inpatient and outpatient domestic treatment, but it does not cover non-medical costs such as transportation to a facility. The program offers treatment options in urban public hospitals, but patients must follow referral procedures. Under CBHI, there is no co-payment and no limit on the number of services provided. However, treatments obtained from private health facilities (except for services that are not available at public facilities) or abroad and treatments which have largely cosmetic value are not covered under the scheme.

In Table 1 we show enrollment, drop-out and new membership rate in CBHI for each region and for various years. The overall enrollment rate was 41% in 2012 but increased to 48% in 2013. 18% of households enrolled in 2012 dropped out and 25% of uninsured households in 2012 became insured in 2013. Enrollment, drop-out and membership renewal varies across regions where pilot districts reside. Scheme uptake seems to be higher in the Amhara region with greater enrollment rates, lower dropout rates and the second highest new membership rate. The lowest enrollment incidence is recorded in SNNP region in both years. Tigray regions show the highest new membership rate at the same time experienced highest dropout rate.

**Table 1:** CBHI enrolment and drop-out in the pilot regions.

| Regions | Enrolled |  | Enrolled |  | Dropped-Out |  | New Members |  |
| --- | --- | --- | --- | --- | --- | --- | --- | --- |
|  | 2012 |  | 2013 |  | 2013 |  | 2013 |  |
|  | % | obs | % | obs | % | obs | % | obs |
| Tigray | 25.65 | 99 | 37.66 | 145 | 26.53 | 26 | 25.44 | 73 |
| Amhara | 37.15 | 146 | 47.07 | 185 | 6.85 | 10 | 19.84 | 49 |
| Oromia | 33.25 | 132 | 33.08 | 132 | 21.21 | 28 | 10.49 | 28 |
| SNNP | 27.3 | 107 | 27.3 | 107 | 21.5 | 23 | 8.07 | 23 |
| Total | 30.87 | 484 | 36.27 | 569 | 18.01 | 87 | 15.93 | 173 |

This table report CBHI take-up, dropout, and new membership in the balanced data.

### Data

In this paper, we used three rounds of household panel data from rural Ethiopia, which was collected as part of the research project on Poverty Dynamics, Health Shocks and Coping Strategies in Ethiopia. This was a collaboration project of the International Institute of Social Studies (ISS) of Erasmus University Rotterdam, the Ethiopian Economics Association (EAA) and the Organization for Social Science Research in Eastern and Southern Africa (OSSREA).

The first round of surveys was collected between March and April in 2011, a few months before the launch of the CBHI program. The subsequent rounds were collected again between March and April in 2012 and between March and April in 2013 after the launch of the CBHI. The data collection involved 8 teams, each comprising 4 enumerators and 1 supervisor. Each team was assigned to collect data from 2 districts.

The surveys cover 16 districts located in four main regional states of Ethiopia, namely Tigray, Amhara, Oromia and SNNP. Twelve of these districts operate CBHI program, while one district in each region did not start offering CBHI. The survey includes a information about individuals, households and community level questions. When collecting the data, 6 villages from each district were selected randomly and 17 households from each village were randomly selected from lists kept by the village administrations. The first round of the survey thus included 1,632 households which were tracked longitudinally. In the second survey round (in 2012) it was possible to resurvey 1,599 households, leading to a 2% attrition. The third survey round contain 1,583 of the households, with an overall 3% attrition from the original sample.

Given that enrollment is accepted at a household rather than the individual level, we are interested in household level lifestyle and timing of treatment seeking measures. Our outcome variables are the following. First, we construct a binary indicator variable that takes a value one if individuals in a household wash hands before and after meals and zero otherwise. Second, another binary indicator variable that takes a value one for households that treat water before drinking (filters, boils or apply other methods to make it safe) and zero otherwise.

In the data we have information about what households would do if symptoms of disease, specifically malaria, tetanus, and tuberculosis, were detected in an adult member of the household. More than 95% of the households stated that they would take them to modern healthcare providers. There is a follow-up question asking when they would take them to their choice of modern care. The answer to this question for Malaria, Tetanus and Tuberculosis (TB) is categorically 1 if they would take them immediately, 2 if they would take them the next day if symptoms persist, 3 if they would take them after two days if symptoms persist, 4 if they would take them to care between three days and a week, 5 if they would take them to care after a week if symptoms persist, and 6 if they would take them to care after more than a week if symptoms persist. Each stage indicates households behavioral response to waiting time for each illness episode, allowing us to investigate the effects of CBHI on whether the insured tends to receive immediate or delayed treatment. These outcomes are based on clinical health need vignettes, where each household responded to hypothetical scenario questions. For more details, refer to Appendix.

Generally, the literature assessing the association between health insurance and lifestyle mainly focuses on individual behavior in relation to diet, smoking, alcohol drinking and exercise, (18,20,22,33). Unfortunately, we do not have this information to extend our analysis in this direction. However, we believe these outcomes would only be marginally relevant for our purposes as we expect lower prevalence of smoking or alcohol consumption and little physical activity due to limited infrastructure in the rural areas where our data were collected.

In our study, the selection of washing hands and treat water or not as key outcomes stems from a clear perspective. While CBHI is traditionally expected to promote health-conscious behaviors, we recognize that in certain contexts, it might lead to riskier behavior due to the financial security it provides. This paradoxical effect may also extend to the timing of healthcare-seeking behavior for illness symptoms. Specifically, while having CBH expected to encourage immediate care-seeking, can, in fact, lead individuals to perceive minor symptoms as less urgent and delay care. Particularly, the reduction in financial barriers can lead some to feel less urgency in seeking care for what they consider minor symptoms. Additionally, the insured may incur additional costs when visiting a healthcare facility, including transportation, lost work time, accommodation, meals, and incidentals. Even though the medical costs are covered, some may perceive their symptoms as not worth the related expense, leading to delayed care.

### Ethics Statement

Ethics approval for the study was obtained from the Ethics Committee at the International Institute of Social Studies, Erasmus University Rotterdam (reference: iss0001946). Committee is chaired by the Deputy Rector for Research Affairs (then, Professor Mohamed Salih).

Our study participants are household heads and we interviewed individuals who are aged above 18 years old and consent regarding child participants is not applicable in our case.

The data collection involved sequential rounds of household recruitment, starting on March 1, 2011, and concluding on April 30, 2011, with subsequent rounds conducted at one-year intervals and two-year intervals at the same time window. Participants (household heads) provided verbal informed consent for their participation in the study. The verbal consent process involved a detailed explanation of the study purpose, procedures, and potential benefits. Participants verbally expressed their willingness to participate, and this process was documented by the interviewer’s team.

### Empirical approach

To examine the impact of CBHI on household lifestyles and timing of healthcare seeking, we implement a household fixed-effect model, exploiting the longitudinal nature of our data. We specify our fixed effects model as follows:

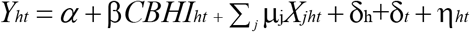

Where *Y*_*ht*_ represents the household-level outcomes at time *t*, which includes handwashing before meals, water treatment before drinking, and the timing of care-seeking for illness symptoms (Malaria, Tetanus, and TB). CBHI is a binary outcome that distinguishes the insured and uninsured households. Hence, β is our parameter of interest, which capture the effect of being insured by CBHI. We include a vector *X*_*jht*_ of covariates that might potentially influence both the outcomes and CBHI enrollment. The controls included are indicator for male household head, household size, number of under 6 years old children, number of male youth (6-15 years), number of female youth (6-15 years), number of working age women (16-64 years), number of elderly (over 64 years old), time to nearest health center, time to nearest hospital, time to all-weather road, households with improved water and modern light sources. We also include education level of the head of household, education level of the spouse, marital status of the head of household, his/her main religion, household consumption per capita, and the region of residence of the household. δ_h_ and δ_*t*_ represent household and time fixed effects, respectively. The household fixed effect allows us to control for any observed or unobserved time invariant characteristics that affect the outcomes and enrollment decisions. And by including time fixed effect, we take into account general changes in lifestyle over the years. η_ht_ denotes the idiosyncratic error term.

To account for potential correlation across households within the same geographic area, we cluster standard errors at the village level (34,35).

Results

### Baseline characteristics of the insured and insured households

Table 2 reports the 2011 baseline characteristics of insured and uninsured households in both pilot and non-pilot woredas. In our sample, both groups are comparable in terms of household composition, but insured households are, on average, farther from health centers, hospitals, and all-weather roads. Access to improved water, and modern light sources is slightly better among the insured.

**Table 2:** Baseline characteristics of control variables between insured and uninsured (balanced panel)

| Independent Var | Insured household (2011) |  | Uninsured household (2011) |  |
| --- | --- | --- | --- | --- |
|  | Mean | Std. Dev. | Mean | Std. Dev. |
| Household composition |  |  |  |  |
| Head sex (Male= 1) | 0.88 | 0.32 | 0.85 | 0.36 |
| Household size | 6.06 | 2.17 | 5.6 | 2.19 |
| Number of under 6 children | 0.14 | 0.15 | 0.16 | 0.17 |
| Number of young male (6-15) | 0.16 | 0.15 | 0.15 | 0.16 |
| Number of young age female (6-15) | 0.15 | 0.14 | 0.14 | 0.15 |
| Number of working age female (16-64) | 0.25 | 0.14 | 0.25 | 0.15 |
| Number of elderly (age>64) | 0.04 | 0.11 | 0.06 | 0.16 |
| Time to health center | 67.46 | 45.04 | 60.58 | 40.67 |
| Time to hospital | 113.57 | 67.31 | 105.87 | 68.74 |
| Time to all weather road | 37.7 | 37.71 | 31.93 | 35.92 |
| Have improved water source | 0.78 | 0.41 | 0.77 | 0.42 |
| Have modern light source | 0.08 | 0.27 | 0.07 | 0.25 |
| Education Status Reference- No education |  |  |  |  |
| Head education informal | 0.16 | 0.37 | 0.09 | 0.29 |
| Head education primary | 0.37 | 0.48 | 0.35 | 0.48 |
| Head education secondary | 0.05 | 0.21 | 0.03 | 0.18 |
| Spouse education informal | 0.08 | 0.27 | 0.03 | 0.17 |
| Spouse education primary | 0.18 | 0.38 | 0.17 | 0.38 |
| Spouse education secondary | 0.02 | 0.13 | 0.02 | 0.12 |
| Marital status Reference-Single |  |  |  |  |
| Married | 0.87 | 0.34 | 0.83 | 0.38 |
| Divorced | 0.05 | 0.22 | 0.06 | 0.24 |
| Windowed/other | 0.07 | 0.26 | 0.1 | 0.3 |
| Religion Reference-Other or no religion |  |  |  |  |
| Orthodox | 0.59 | 0.49 | 0.43 | 0.49 |
| Protestant | 0.14 | 0.35 | 0.26 | 0.44 |
| Muslim | 0.27 | 0.44 | 0.27 | 0.44 |
| Per capita consumption Reference- |  |  |  |  |
| Quintile 2 | 0.18 | 0.39 | 0.23 | 0.42 |
| Quintile 3 | 0.21 | 0.41 | 0.19 | 0.39 |
| Quintile 4 | 0.22 | 0.42 | 0.18 | 0.39 |
| Quintile 5 | 0.21 | 0.4 | 0.18 | 0.39 |
| Region Reference Tigray |  |  |  |  |
| Amhara | 0.33 | 0.47 | 0.15 | 0.36 |
| Oromia | 0.26 | 0.44 | 0.24 | 0.43 |
| SNNP | 0.18 | 0.38 | 0.34 | 0.47 |
| Total observations | 862 |  | 708 |  |

The insured households have better-educated heads and spouses. The heads are more married, less divorced, and widowed compared to uninsured household heads. In terms of religious denomination, most of the heads of the insured are Orthodox Christians, while Muslims are relatively dominant among the uninsured. On average, per capita consumption is higher for insured households. The majority of the insured group in our sample originates from Amhara, which is Ethiopia’s second most populous region. In contrast, a higher proportion of uninsured households are located in the SNNP region.

### Trends in lifestyle measures and healthcare seeking behavior

In Table 3 we show the average level of each outcome by treatment status (insured versus uninsured households - pilot and non-pilot) and survey year. The statistics prior to CBHI implementation is based on enrollment status in post CBHI intervention period (2012, and 2013). Washing hands before meals shows a slight increase in insured households over the years, while this trend decreased in 2013 among uninsured households. When it comes to water treatment, the trend is more V-shaped among the insured but reverses among the uninsured. Among the insured, the preferred waiting time for treatment of Malaria, Tetanus, and TB appears to have been dropped immediately after CBHI launch, but then increased in 2013. While for the uninsured, with the exception of tetanus, the trend is consistently downward.

**Table 3:** Trends in household lifestyle indicators by enrollment status (balanced panel)

| Dep. var | Insured Households |  |  |  |  |  | Uninsured Households |  |  |  |  |  |
| --- | --- | --- | --- | --- | --- | --- | --- | --- | --- | --- | --- | --- |
|  | 2011 |  | 2012 |  | 2013 |  | 2011 |  | 2012 |  | 2013 |  |
|  | Mean | Std. Dev. | Mean | Std. Dev. | Mean | Std. Dev. | Mean | Std. Dev. | Mean | Std. Dev. | Mean | Std. Dev. |
| Wash hands | 0.95 | 0.21 | 0.97 | 0.16 | 0.99 | 0.08 | 0.88 | 0.33 | 0.93 | 0.25 | 0.94 | 0.23 |
| Water treatment | 0.18 | 0.38 | 0.17 | 0.38 | 0.07 | 0.26 | 0.10 | 0.30 | 0.14 | 0.34 | 0.07 | 0.25 |
| Seek care malaria | 2.23 | 1.12 | 2.30 | 1.04 | 2.12 | 1.14 | 2.59 | 1.35 | 2.31 | 1.01 | 2.24 | 1.16 |
| Seek care tetanus | 2.19 | 1.32 | 2.03 | 1.14 | 2.23 | 1.36 | 2.68 | 1.51 | 2.20 | 1.12 | 2.29 | 1.31 |
| Seek care TB | 2.76 | 1.53 | 2.72 | 1.39 | 2.79 | 1.70 | 3.32 | 1.72 | 2.80 | 1.31 | 2.74 | 1.61 |

### Baseline estimates

In Table, 4 we present the effects of CBHI on household lifestyle indicators. We offer these estimates in two distinct ways: First, by comparing insured households with uninsured households in the pilot districts. Second, by combining uninsured households from both pilot and non-pilot districts as the control group. This approach allows us to assess how the effects vary depending on the composition of the control households, providing insights into the sensitivity and generalizability of the findings.

**Table 4:**
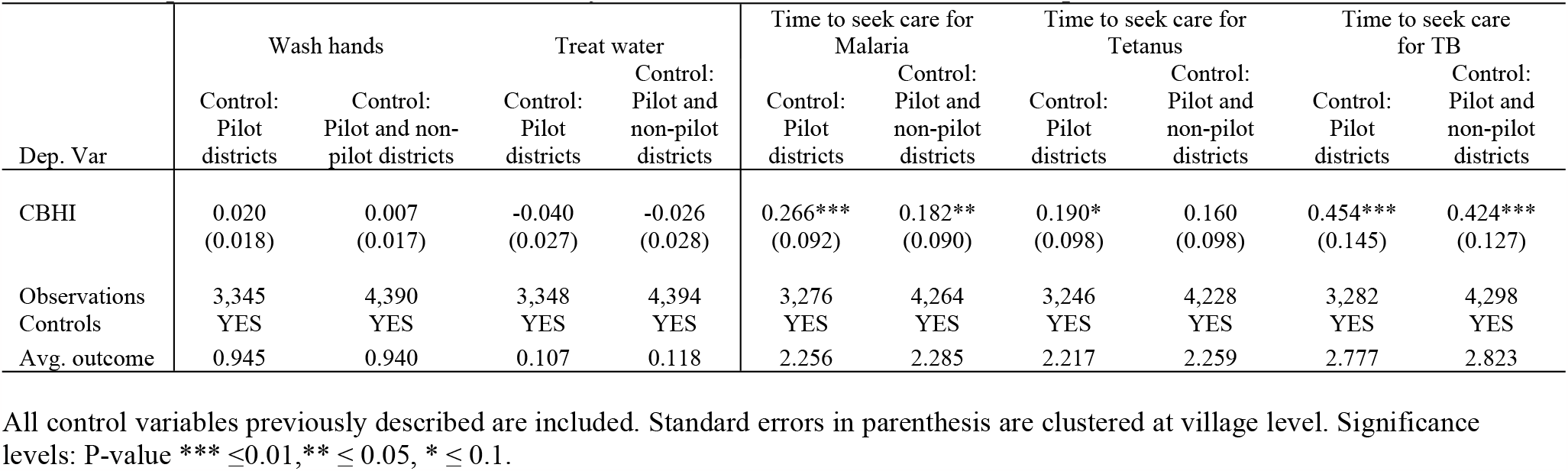
Impact of CBHI on household lifestyle: Fixed effect estimates (balanced panel)

| Dep. Var | Wash hands |  | Treat water |  | Time to seek care for Malaria |  | Time to seek care for Tetanus |  | Time to seek care for TB |  |
| --- | --- | --- | --- | --- | --- | --- | --- | --- | --- | --- |
|  | Control: Pilot districts | Control: Pilot and non-pilot districts | Control: Pilot districts | Control: Pilot and non-pilot districts | Control: Pilot districts | Control: Pilot and non-pilot districts | Control: Pilot districts | Control: Pilot and non-pilot districts | Control: Pilot districts | Control: Pilot and non-pilot districts |
| CBHI | 0.020<br>(0.018) | 0.007<br>(0.017) | -0.040<br>(0.027) | -0.026<br>(0.028) | 0.266***<br>(0.092) | 0.182**<br>(0.090) | 0.190*<br>(0.098) | 0.160<br>(0.098) | 0.454***<br>(0.145) | 0.424***<br>(0.127) |
| Observations | 3,345 | 4,390 | 3,348 | 4,394 | 3,276 | 4,264 | 3,246 | 4,228 | 3,282 | 4,298 |
| Controls | YES | YES | YES | YES | YES | YES | YES | YES | YES | YES |
| Avg. outcome | 0.945 | 0.940 | 0.107 | 0.118 | 2.256 | 2.285 | 2.217 | 2.259 | 2.777 | 2.823 |

All control variables previously described are included. Standard errors in parenthesis are clustered at village level. Significance levels: P-value \*\*\* $\leq 0.01$ , \*\* $\leq 0.05$ , \* $\leq 0.1$ .

Our estimate shows that participation in the CBHI scheme does not affect behavior related to hygiene and water treatment before drinking regardless of the location of control households. However, in terms of waiting time, we find a delay of approximately 0.18 to 0.27 points in the average treatment seeking timing outcome for Malaria symptoms in insured households compared to uninsured households. Given the scale of the dependent variable where higher levels indicate longer delays, this suggests that having CBHI is associated with a modest increase in delaying the timing of treatment. This result is statistically significant irrespective of the composition of control households, but the point estimates slightly decreases when this composition is both from the pilot and non-pilot districts. There is also a significant but relatively weak effect on preferred waiting time for Tetanus treatment. In this case, CBHI participation results in 0.19 point increases in waiting time for tetanus treatment when comparing the insured versus the uninsured in the pilot districts only. This marginal effect disappears when we consider the uninsured households from both pilot and non-pilot districts. Finally, participation in CBHI also increase relatively substantial delay (0.45 - 0.42 points more in the average treatment timing) in treatment timing if a TB illness symptoms are seen in adult member of the household.

### Robustness

#### Dichotomizing timing of treatment seeking behavior

We assess the sensitivity of our baseline estimates by converting treatment seeking outcome variables into binary indicators, measuring the effect on extensive margin. Specifically, we create an outcome that takes a value of one if a household would seek care immediately or the next day and zero otherwise for each illness symptoms (malaria, tetanus, and tuberculosis). Similar to the baseline, we assess the impact in a setting where the control groups are initially the uninsured in pilot districts and where the controls consist of uninsured households in pilot and non-pilot districts.

Table 5 shows that our findings are robust, with the expectation of tetnus. In particular, we find a reduction in immediate or next-day medical care seeking behavior among insureds compared to uninsured in pilot districts or uninsured in both pilot and non-pilot districts. In fact, the effect on tetanus is also robust, since initially the effect on tetanus symptoms was weaker. In overall, the estimates confirms that, as compared to those who are uninsured, the incidence of seeking immediate medical care for insured individuals are less by 7-10 percentage points for malaria and by 7-7.4 percentage points for tuberculosis.

**Table 5:**
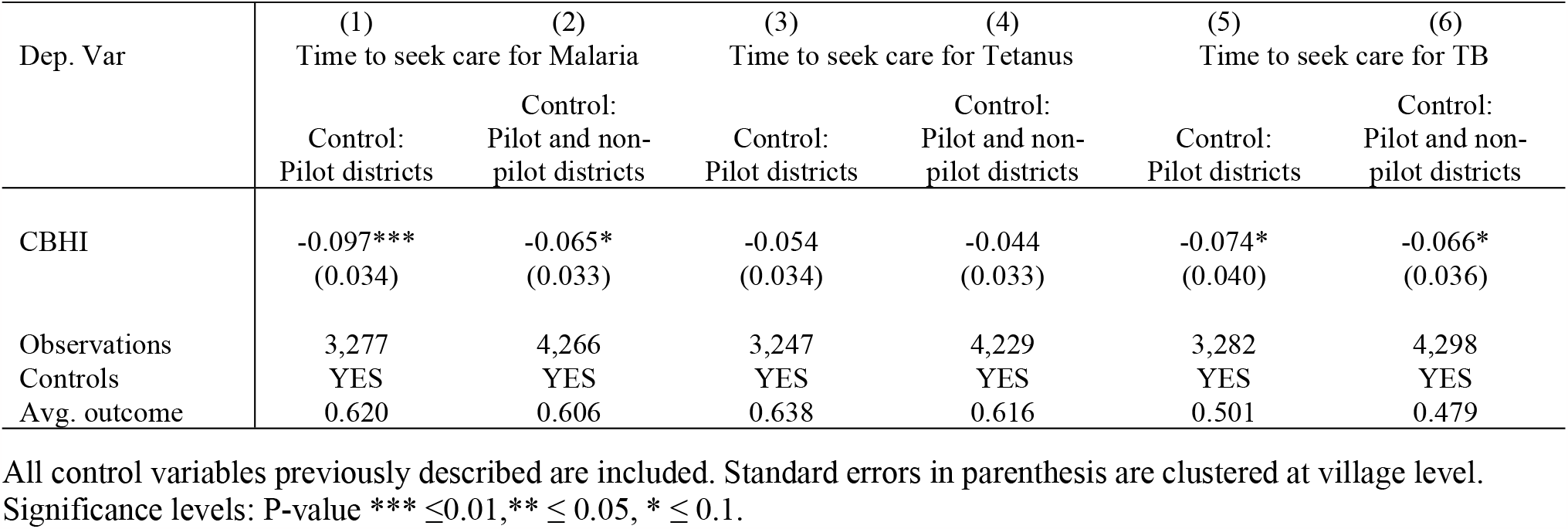
Impact of CBHI on treatment seeking behavior: Extensive margin (balanced panel)

#### Fixed effect Ordered logit estimator

Our baseline findings, obtained through a linear fixed effect estimator, are based on the assumption of equidistant intervals between the six categories of the timing of treatment-seeking outcome. We explore the sensitivity of our results to this assumption by employing a fixed effect ordered logit model with blow-up and cluster estimator. Unlike the linear model, the ordered logit estimator only assumes the ordered nature of response options and does not presuppose equal intervals between them (26). We adopt feologit user written command contributed by (36).

The results in Table 6 is generally in agreement with the baseline finding. Particularly, insured households exhibit 1.36 to 1.61 times higher odds of falling into a higher wait time category for malaria symptom treatment and 1.70 to 1.77 times higher odds for tuberculosis treatment compared to uninsured households, with all other model variables held constant. Similar to the baseline, this estimator shows a weak effect in the case of tetanus symptoms. Overall, this estimator appears to sustain the baseline finding that enrollment in CBHI is associated with delay in treatment.

**Table 6:** Impact of CBHI on the timing of treatment seeking behavior: Ordered logit regression.

| Dep. Var | (1) | (2) | (3) | (4) | (5) | (6) |
| --- | --- | --- | --- | --- | --- | --- |
|  | Time to seek care for Malaria |  | Time to seek care for Tetanus |  | Time to seek care for TB |  |
|  | Control:<br>Pilot districts | Control:<br>Pilot and non-pilot districts | Control:<br>Pilot districts | Control:<br>Pilot and non-pilot districts | Control:<br>Pilot districts | Control:<br>Pilot and non-pilot districts |
| CBHI | 1.608***<br>(0.260) | 1.359**<br>(0.208) | 1.336*<br>(0.201) | 1.281*<br>(0.192) | 1.772***<br>(0.317) | 1.703***<br>(0.269) |
| Observations | 2869 | 3692 | 2792 | 3565 | 2978 | 3872 |
| Controls | YES | YES | YES | YES | YES | YES |
| Avg. outcome | 2.288 | 2.326 | 2.292 | 2.352 | 2.829 | 2.875 |

This table reports the odds ratios from a fixed effect ordered logit estimator. Standard errors in parenthesis are clustered at village level. Significance levels: P-value \*\*\* $\leq 0.01$ , \*\* $\leq 0.05$ , \* $\leq 0.1$ .

## Discussion

Overall, while we don’t find changes in preventive care activities, CBHI appears to increase delay in seeking care for illness symptoms. Delayed use of primary health care use could increase the risk of developing worse health conditions and may substantially increase health care costs. It could also create considerable challenges for the financial sustainability of insurance schemes (14,37,38). Since the Ethiopian CBHI is primarily intended for rural residents who are largely poor, an immediate visit for symptoms of illness would be costly to them. This is because, in rural areas, mainly health posts are available, but these facilities engaged in provision of preventive and promotion health services, and they do not provide curative care. In order to get medical care, they need to travel to the urban areas where health centers and hospitals are established. Visiting these facilities involves transportation infrastructure and subsistence costs that are not covered by CBHI scheme.

The existing evidence also indicates that one of the reasons for low utilization of modern care in rural Ethiopia is transportation barriers (39,40). In addition, seeking immediate care for rural people would result in disruption of productive activity and loss of income while off farming. This opportunity cost might weigh more than the benefit from the household’s perspective (41). This rationale aligns with evidence from rural China, where factors such as involvement in farming, longer distances to the hospital, and awareness were significantly associated with higher odds of delaying care for TB (42). Knowledge is likewise reported as the key factor in delaying TB treatment in Ethiopian context (43,44) and for malaria higher waiting time at the health facility was associated with increased delay in treatment (45). However, our finding may contrast (46) which reveals the reverse, highlighting delayed treatment-seeking for actual TB symptoms among individuals without health insurance.

In the context of preventive care, our findings aligns with studies by (24,47–49), which also report insignificant impacts of other health insurance schemes. However, the broader literature paints a varied picture, with the majority of studies showing significant effects that depend on the scope of insurance coverage and the country of study. For instance, (28) reported significant increase in unhealthy practices related to the use of malaria protective materials among members of the Ghanaian National Health Insurance Scheme, although this finding contradicts the results of a randomized experiment (26). In a similar insurance context, (20) suggested that participation in China’s Rural Cooperative Medical Scheme encourages risky behaviors, such as smoking, heavy drinking, low physical activity, consumption of high-calorie food, and being overweight. Studies in countries with advanced insurance programs, (21,22,50), have shown increases in unhealthy individual behaviors, such as heavy smoking, weight gain and lack of exercise in relation to private insurance and Medicare. In contrast, (18,23,33) indicates that participation in insurance programs can improve preventive care activities. This disparity is likely due to variations in outcomes, research methodologies, and contextual factors.

In (19) a distinction is made between various sources of insurance to identify which ones lead to increased risky behavior. The findings indicate that public health insurance may contribute to higher inefficiencies in terms of exacerbating unhealthy behavior when compared to private insurance. This suggests that, as observed in the Ethiopian context, public health insurance schemes may face challenges in effectively controlling unhealthy practices such as delay in care seeking among insurance beneficiaries and in developing efficient insurance programs.

The results of this work shed new light on the understanding of the impact of CBHI scheme beyond its primary objectives of promoting healthcare utilization and reducing the burden of out-of-pocket health expenditure. While many CBHI schemes in developing country aim to counteract adverse selection, little attention has been paid to possible adverse behavioral responses from members. The evidence in this study implies the need to identify and solve major causes of delay in seeking medical services among health insurance members. This is important step to promote early use of services and, consequently, maintain financial sustainability of the Ethiopian CBHI scheme.

## Conclusion

There is an increasing number of studies highlighting concern about the financial sustainability of CBHI schemes in developing countries. We partially respond to such concern by examining the potential contribution of lifestyle and delayed care seeking behavior in the context of Ethiopian CBHI. We find that CBHI increased waiting time for treatment for malaria, tetanus and tuberculosis symptoms. The key important implication for the insurer is that this delayed treatment would incur higher costs than immediate treatment. If there are many such insured households delaying treatment, perhaps the CBHI could collapse due to a large claim for reimbursement of care costs.

The expansion of CBHI in conjunction with supply-side investments such as improving the accessibility of healthcare could mitigate such behavioral responses from members. Also, improving the quality of care and increasing the staff could potentially reduce waiting times at the health center, which could partially reduce the opportunity cost of seeking immediate care.

Due to data limitation, we could not further explore behavioral responses in terms of other indicators such as physical exercise, smoking, and alcohol consumption and potential underlying mechanisms for the reported results. There is, therefore, ample scope for future research in this area, including our limitations. CBHI is often considered a better alternative to out-of-pocket expenditure on the path towards achieving universal health coverage. Therefore, finding out other dimensions of factors that contribute to counteracting CBHI in developing country is very important.

## Data Availability

Due to data protection restrictions within collaborative research organizations, we are currently unable to share the data. Regarding the condition of replicating our results, we can share both the data and the do-file upon reasonable request. For this purpose, please contact Dr. Alemu at or call +251911529796

## Appendix

### Health Care Need Vignettes

Respondents were asked what they would do if they or someone in their household-were to experience the following symptoms themselves. Only one answer per question was allowed. Interviewers were instructed to emphasize that respondents should state what they would do – not what they think should be done. The questions are:

A 20 year old male has always been healthy. For the last week, he has episodes of sudden coldness followed by rigor and then fever and sweating. These episodes occur about every two days. In between episodes he can still do some light housework.

A 25 year old male has got a small cut in his leg when working on the field three days ago. The wound has become red and from time to time he feels a throbbing pain in his leg, but he can still walk around and do some work.

A 35-year-old female has been coughing for three weeks now. She feels more tired than usual but can still do some housework. Her relatives think she looks thinner than a few weeks ago.

The answers to when would you go to facility of choice?

1=immediately,

2=the next day if symptoms continue, 3=after two days if symptoms continue,

4=between three days and a week if symptoms continue, 5=after a week if symptoms continue,

6=after more than a week if symptoms continue

